# ChatGPT in glioma patient adjuvant therapy decision making: ready to assume the role of a doctor in the tumour board?

**DOI:** 10.1101/2023.03.19.23287452

**Authors:** Julien Haemmerli, Lukas Sveikata, Aria Nouri, Adrien May, Kristof Egervari, Christian Freyschlag, Johannes A. Lobrinus, Denis Migliorini, Shahan Momjian, Nicolae Sanda, Karl Schaller, Sebastien Tran, Jacky Yeung, Philippe Bijlenga

**Author notes:** Julien Haemmerli and Lukas Sveikata contributed equally to this work. Corresponding author: Julien Haemmerli, MD, Division of Neurosurgery, Department of Clinical Neurosciences, Geneva University Hospitals and Faculty of Medicine, University of Geneva, Rue Gabrielle-Perret-Gentil 4, 1205 Geneva, Switzerland.

## Abstract

**Introduction:** ChatGPT, a novel AI-based chatbot, sparked a lot of interest in the scientific community. Complex central CNS tumour cases require multidisciplinary expert recommendations that incorporate multimodal disease information. Thus, the potential of ChatGPT to integrate comprehensive treatment information may be of tremendous benefit for CNS tumour decision-making. We evaluated the ChatGPT recommendations for glioma management by a panel of CNS tumour experts.

**Methods:** We randomly selected 10 patients with primary CNS gliomas discussed at our institution’s Tumour Board. Patients’ clinical status, surgical, imaging, and immuno-pathology-related information was provided to ChatGPT and seven CNS tumour experts. The chatbot was asked to give the most likely diagnosis, the adjuvant treatment choice, and the regimen while considering the patient’s functional status. The experts rated the AI-based recommendations from 0 (complete disagreement) to 10 (complete agreement). An intraclass correlation agreement (ICC) was used to measure the inter-rater agreement.

**Results:** Eight patients (80%) met the criteria for glioblastoma and two (20%) were low-grade gliomas. The experts rated the quality of ChatGPT recommendations as poor for diagnosis (median 3, IQR 1-7.8, ICC 0.9, 95% CI 0.7-1.0), good for treatment recommendation (7, IQR 6-8, ICC 0.8, 95% CI 0.4-0.9), good for therapy regimen (7, IQR 4-8, ICC 0.8, 95% CI 0.5-0.9), moderate for functional status consideration (6, IQR 1-7, ICC 0.7, 95% CI 0.3-0.9), and moderate for overall agreement with the recommendations (5, IQR 3-7, ICC 0.7, 95% CI 0.3-0.9). No difference were observed between the glioblastomas and low-grade glioma ratings.

**Conclusions:** ChatGPT performed poorly in classifying glioma types but was good for adjuvant treatment recommendations as evaluated by CNS Tumour Board experts. Even though the ChatGPT lacks the precision to replace expert opinion, it may become a promising tool to supplement experts, especially in low-resource settings.

## INTRODUCTION

Artificial intelligence (AI) is attracting a lot of interest in the present era of personalized medicine. ^1–3^. Since novel drug discovery, surgical robotics or complex interdisciplinary oncologic therapy decisions are time-consuming and resource demanding, innovative AI-based language models are being developed with the aim of boosting healthcare professionals’, scientists’, and bioengineers’ performance ^4–6^. Recently, a novel AI-based chatbot, ChatGPT, was launched, spurring the curiosity and the skepticism of the scientific community ^7–11^. ChatGPT is based on a deep learning model called the *Generative Pre-trained Transformer* (GPT). This results in a language model that uses unsupervised learning to generate human-like text ^12^. In its interface, the user is able to chat with the underlying AI as one would interact with an expert.

Neurooncolgy has significantly evolved in parallel with new research advances ^13^. For instance, treatment of high-grade gliomas has been extensively studied for the last 20 years to offer a longer survival rate for affected individuals ^14,15^. Furthermore, the consideration of the patient’s clinical state, age and comorbidities have been included in novel trials to optimize treatment protocols ^16^. Low-grade gliomas account for approximately 20% of all gliomas. Their management is more heterogenous and the adjuvant treatment is based on the complex molecular profile ^17–19^. In order to determine the best adjuvant treatment for glioma patients, central nervous system (CNS) tumour boards (TB) arose implicating a multidisciplinary team composed of neurosurgeons, oncologists, neurologists, pathologists, radiation oncologists and neuroradiologists ^20^. TBs are, however, mobilizing a vast amount of resources, which might be challenging to apply for every patient suffering from a glioma. In this regard, AI-assisted decision-making could prove helpful in delivering personalized treatment strategies ^21^.

Given the promise of AI in utilizing vast sources of information to synthesize information and provide recommendations, we investigated whether ChatGPT had a role to play in CNS TB regarding glioma patient adjuvant therapy decision-making. We hypothesized that ChatGPT’s performance was at least as good as a CNS TB to subclassify gliomas ^22^ and to propose an adjuvant therapy in line with the current guidelines.

## METHODS

### Patients selection

We randomly selected 10 glioma cases from our institutional CNS TB registry from 2014 and 2022. During this period a total of 215 cerebral glioma cases were evaluated. Inclusion criteria were: 1) new onset or recurrent supratentorial glioma, 2) surgical treatment was performed (removal or biopsy) 3) CNS TB recommendation, and 4) informed consent was available. Exclusion criteria were: 1) a presence of brain metastasis 2) extra-axial tumours, and 3) glioma involving the brainstem or the spinal cord.

### Dialogue with ChatGPT

Electronic patients’ records were retrospectively reviewed. From the first of February to the 14^th^ of February 2023, case summaries were presented to ChatGPT, as it would be presented at our institutional CNS TB. The description contained the main clinical information, context of admission, preoperative radiological and surgical information, postoperative clinical information, neuropathological findings, and results of the immunohistochemical and molecular examination. No diagnosis nor patient identification information was provided to ChatGPT. Two questions were then asked to ChatGPT: 1) “What is the best adjuvant treatment?”, 2) “What would be the regimen of radiotherapy and chemotherapy for this patient?”. ChatGPT’s answers were collected. A new chat box was opened for each case, in order to avoid learning from the same conversation. A complete chat transcript, including case description and ChatGPT’s output are provided in the supplementary material.

### CNS Tumour Board, and experts selection

Our institutional CNS TB is composed of neuro-oncologists, radio-oncologists, radiologists, neurosurgeons, neuropathologists, and neurologists. We considered our institutional CNS TB as a reference, as its decisions are evidence-based and are supported by a multidisciplinary consortium. In our institution, every patient with CNS oncological disease is presented at this multidisciplinary meeting. For the purpose of this study, five experts from our CNS TB (two neuropathologists, one neurosurgeon, one radio-oncologist, and one oncologist) and two external independent experts (two neurosurgeons from Europe and the United States) evaluated ChatGPT’s output with regard to the formal decision of the CNS TB.

### Studied parameters

The experts were asked to rank ChatGPT’s answers for each of the ten cases. The CNS TB decisions were used as the gold standard. The experts were asked to evaluate the ChatGPT’s output on a scale between 0 and 10, where “0” indicated complete disagreement, 10 indicated complete agreement, and 5 a neutral answer (“neither agreement nor disagreement”). The experts had to evaluate ChatGPT’s answers regarding the diagnosis, the proposed treatment, the consideration of the patient’s functional status to support adjuvant therapy, the proposed regimen of adjuvant therapy, and the overall accuracy of ChatGPT with respect to its answers. Finally, the experts were asked to provide their opinion on the possible place of AI in interdisciplinary CNS tumour decision-making. The experts were provided with a questionnaire to rate ChatGPT’s performance in providing the diagnosis of specific glioma types, adjuvant treatment recommendations, adjuvant therapy regimen, how well the chatbot integrated the overall functional status of the patient into the decision-making, and what is the overall quality of the recommendations provided (supplementary material). Figure 1 summarizes the methods of this study. Supplementary material 2 presents the questions asked to the experts. Finally, the agreement between experts was evaluated.

**Figure 1.**
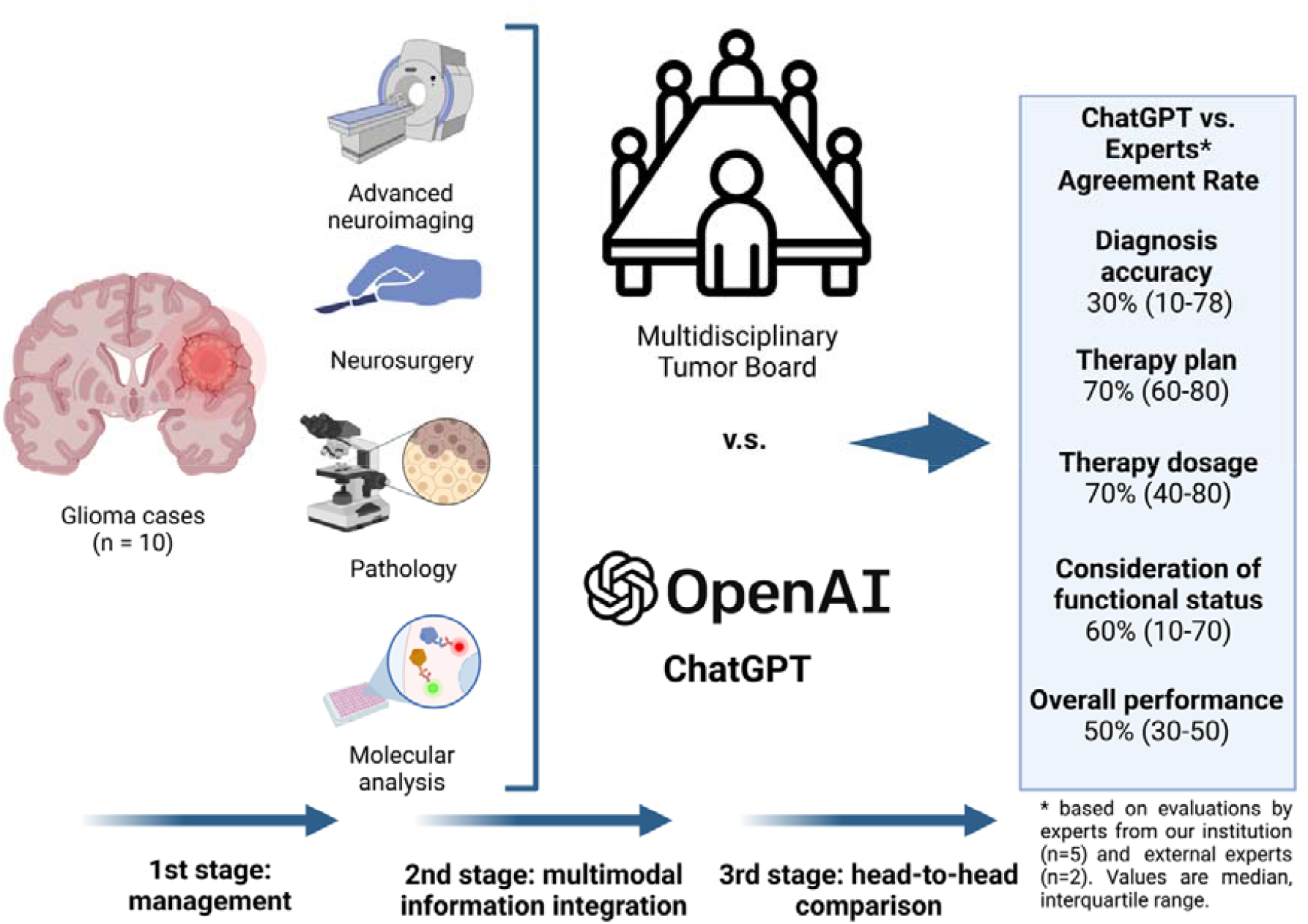
Summary of study workflow. Ten patients were randomly selected from our institutional central nervous system (CNS) tumour board (TB) registry and included in the study. Second, all participants received state-of-the-art pre- and postoperative glioma workup. Third, a summary of the anonymized case and immunohistological findings were presented to the ChatGPT, as it would be done at the CNS TB. Seven experts compared ChatGPT’s output and the TB recommendations. The results represent the median experts’ rating with the interquartile range. The figure was created with BioRender.com.

**Figure 2.**
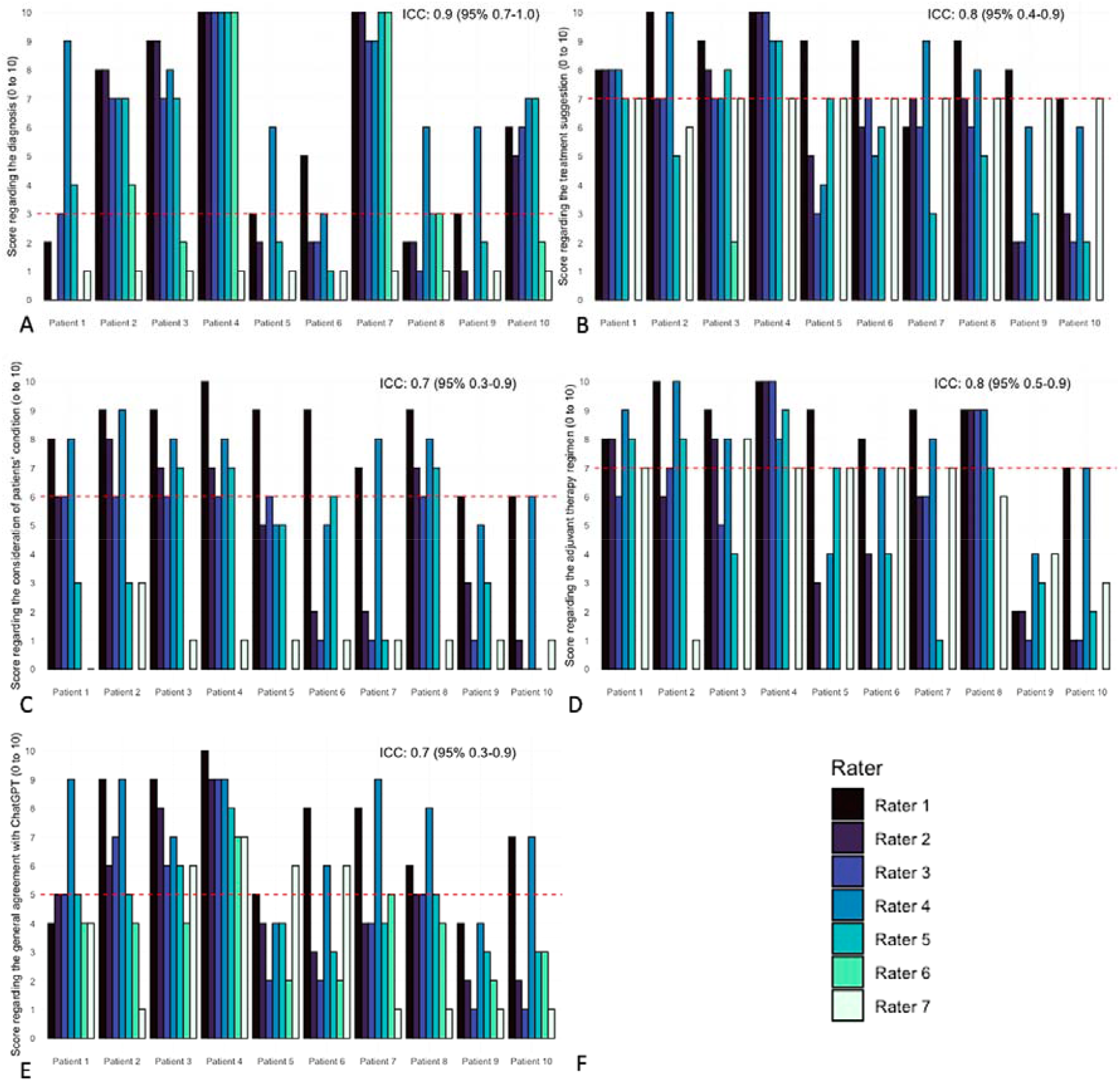
Barplots representing the ratings per patient and per expert, regarding (A) the diagnosis, (B) the adjuvant treatment recommendation, (C) the consideration of the patient’s functional status, (D) the regimen of the adjuvant therapy, (E) ChatGPT’s overall performance, (F) the legend. ICC: intraclass correlation coefficient (from 0 to 10, 95% CI). The dashed red line represents the median value of the experts’ rating.

### Statistics

We used R version 3.6.1 for the statistical analysis. The randomization process was performed using function floor(runif). Ordinal variables were presented as median with interquartile range and were compared using a Mann-Whitney U-test when appropriate. Experts’ rating score between 0 to 3 was considered poor, 4 to 6 as moderate, 7 to 8 as good, and 9 to 10 as excellent. The intraclass correlation coefficient (ICC) was used to evaluate the agreement between the experts (two-way random effects, absolute agreement, multiple raters average, ICC (2,*k*)) ^23^. An ICC<0.5 was considered as poor, ≥0.5 and <0.75 as moderate, ≥ 0.75 and <0.9 as good, and ≥ 0.9 as excellent agreement ^23^. Hypothesis testing was considered significant at p-value <0.05 (two-sided).

## RESULTS

### ChatGPT’s output

ChatGPT provided spontaneously the diagnosis for suspected glioma type, recommendations for adjuvant treatment plan, regimen for radio- and chemotherapy, and consideration of functional status for all 10 cases. Regarding the first question “what is the best adjuvant treatment”, ChatGPT started the dialogue by giving its appreciation of the diagnosis. Based on the patient summary, it correctly recognized and classified the tumours as glioma in all cases and suggested the tumour type (e.g. low-grade glioma, grade II or III astrocytoma, glioblastoma). Of note, no alternative diagnosis such as brain metastasis or extra-axial brain tumour was proposed. ChatGPT then recommended “the best adjuvant treatment […]” or “the standard of care for glioblastoma […]”. Concerning the second question “what would the regimen of radiotherapy and chemotherapy be for this patient”, ChatGPT provided a recommendation for all cases. However, complete regimen of radiotherapy (grays in fractions over weeks) was provided in 70% of the cases, and a complete regimen of chemotherapy (medication and doses) in 50% of cases.

For both questions, ChatGPT nuanced its answers for all cases by mentioning the need to adjust the treatment according to the patient’s individual preferences and functional status, although never specifying alternatives. Finally, ChatGPT mentioned the need to confirm its treatment suggestion with a multidisciplinary team in 80% of the cases.

### Experts’ opinion and agreement

Seven experts rated ChatGPT’s output regarding the diagnosis, recommendations for therapy and regimen, and overall accuracy. Rater 6 only rated the diagnosis accuracy and treatment recommendations for case 2 and did not rate the output regarding the consideration of the functional status nor the regimen of adjuvant therapy (expert who preferred to remain in their scope of practice).

Concerning the diagnosis, ChatGPT’s output was evaluated as poor with a median score of 3 (IQR 1-7.8) with excellent agreement between the experts (ICC 0.9, 95% CI 0.7-1.0). For the adjuvant therapy, the ChatGPT recommendations were evaluated as good with a median score of 7 (IQR 6-8) and a good agreement (ICC 0.8, 95% CI 0.4-0.9). The adjuvant therapy regimen was evaluated as good with a median score of 7 (IQR 4-8) and good expert agreement (ICC 0.8, 95% CI 0.5-0.9). Regarding ChatGPT’s output on the consideration of the patient’s functional status, the experts rated the recommendations as moderate with a median score of 6 (IQR 1-7) and a moderate agreement (ICC 0.7, 95% CI 0.3-0.9). Finally, the global evaluation of ChatGPT’s output accuracy was moderate and scored 5 (IQR 3-5) with a moderate expert agreement (ICC 0.7, 95% CI 0.3-0.9). Six experts (86%) evaluated ChatGPT’s role in a CNS TB as useful if the AI-based system can evolve and learn. One rater (14%) evaluated ChatGPT’s role in a CNS TB as useful, but in some specific circumstance.

There was no significant difference between experts’ ratings in glioblastoma (8/10) and low-grade glioma cases.

## DISCUSSION

In this study, we assessed the performance of ChatGPT, an AI-based language generator, in providing treatment recommendations for glioma patients. To the best of our knowledge, this is the first study aiming to evaluate this novel chatbot within the framework of medical multidisciplinary decision-making, such as a CNS TB. While ChatGPT demonstrated proficiency in accurately identifying cases as gliomas, it displayed limited precision in identifying specific tumour subtypes. Furthermore, the tool’s recommendations regarding treatment strategy and regimen were rated as good, while the ability to incorporate functional status in its decision-making process as moderate.

### CNS Tumour board and limitations

Oncologic patients discussed in the multidisciplinary CNS TB are more likely to benefit from a pre- and postoperative staging and are more likely to receive the optimal adjuvant treatment ^24,25^. Barbaro et al. presented in their review the foundations of neuro-oncology and the need for multidisciplinary expertise in order to embrace the multiple disease aspects in CNS tumour-affected patients ^13^. The authors highlighted the prerogatives and missions of a CNS TB: 1) Neuro-oncology, Neurosurgery, Radiation Oncology, Neuropathology, Neurology and Radiology are specialties necessary to compose the CNS TB. 2) The experts consortium’s main goal is to propose a collaborative treatment plan. 3) The development of novel clinical trials. Furthermore, a single-center prospective evaluation of a CNS TB showed that the experts’ consortium influences the clinical management of patients suffering from a brain tumour through high-impact decisions ^26^. However, the organization of regular CNS TB encounters is limited by a number of variables, such as the economic costs, time expenditure, resource availability, and the limited presence of TB across the geographic and socio-economic strata ^25^. New AI-based tools with underlying deep learning, such as ChatGPT, might represent a valuable complement or at least some help to centers lacking expertise or financial resources.

### ChatGPT ready to assume the role of the doctor?

The evaluation of ChatGPT’s recommendations was somewhat varied. On the one hand, ChatGPT was evaluated poorly regarding the diagnosis of the glioma subtype. The output given by the chatbot was often incorrect (i.e., pleiomorphic astrocytoma instead of glioblastoma in case 7), or not detailed enough (i.e. grade II or III astrocytoma in case 4). On the other hand, the adjuvant treatment suggestion and its regimen were rated as good. In this cohort, 80% of the included patients were diagnosed with glioblastoma (WHO grade IV). In the literature, the treatment of glioblastoma WHO IV has been extensively studied ^27–32^. AI models used by ChatGPT are trained on a large dataset of information found online including websites, journals, and digitalized books. It is thus comprehensible that ChatGPT’s output given regarding the adjuvant treatment and its regimen is of good quality because the underlying knowledge base is well-documented.

To this extent, ChatGPT’s performance is mediocre regarding recommendations that are based on less extensive knowledge base. The consideration of patient functional status was rated as moderate, even though the clinical pre- and postoperative state of the included cases was presented to ChatGPT. This consideration is much less documented in the literature as only a few clinical trials studied adjuvant therapy for glioblastoma in impaired functional conditions or in older patients ^30^.

Finally, all ChatGPT’s recommendations were conscientiously mitigated with disclosure statements that medical professionals should validate its suggestions. Furthermore, the chatbot highlighted for several patients that their functional condition is to be considered. Accordingly, and rightfully, the authors suspect that the protocol underlying ChatGPT incorporates restrictions, and its output has been limited regarding providing medical advice. Given this, the authors cannot fully appreciate the full potential of ChatGPT in CNS TB. Notwithstanding this limitation, one could imagine that ChatGPT, with pursued development in this direction, could hold great promise to complement the classic CNS TB workflow.

### Further developments

Six of the seven experts evaluated ChatGPT as useful if the system could learn and improve. This notion is supported by the medical community as AI is growing and holds immense promise in medicine ^2,6,33–35^. However, since its launch in November 2022, ChatGPT has raised skepticism in the scientific community regarding threats to the originality of scientific work ^10,11,36–39^. In particular, there is a lot of concern regarding the possibility that chatbots can be used to write scientific material ^8^. Such concerns have resulted in the development of mitigation strategies, such as AI-generated text detectors ^40^. Similar algorithms aimed at detecting AI-generated text are being developed and implemented by major publishers ^38^. Another consideration is the risk that AI chatbots may be prone to bias or commit omissions and errors in the interpretation of medical information. In the early stages of adoption, these technologies should be used with a human-in-the-loop approach.

Even if our results suggest a reserved rating for ChatGPT’s performance on glioma subtype diagnosis and multi-modal information integration, AI-based chatbots may be a promising supplement in TB decision-making. Future studies could explore ways to refine ChatGPT’s functionality, such as incorporating more patient-specific data and refining its ability to provide nuanced recommendations based on the clinical context. Furthermore, future developments in the ChatGPT interface could introduce the ability to read medical imaging, such as pre- and postoperative brain MRI, which could enormously improve its diagnostic ability and treatment recommendations.

Nonetheless, our results highlight the potential utility of ChatGPT in facilitating clinical decision-making. Chatbots could be used to quickly provide information related to a patient’s medical history, differential diagnosis, relevant diagnostic tests, experimental treatment options, and potential side effects. Furthermore, we intentionally provided the chatbot with only one conversation log. Thus, it is possible that further interaction and additional discussion with the chatbot may have yielded increased performance.

On the other hand, we were somewhat surprised that ChatGPT’s output did not include information on novel clinical trials in the glioma field, which should be accessible to the chatbot algorithm online ^41^. One explanation could be paywall barriers to some of the prominent journals. Thus, breaking these barriers or facilitating AI access to the newest scientific information, could be one potential direction of future development as the new ongoing trials consist of a crucial part of a CNS TB discussion ^13^. With deep learning, chatbots such as ChatGPT could have the potential to integrate the newest trial and bench science information into multidisciplinary decision-making and help TB direct patients to potential applicable clinical trials.

## CONCLUSION

We have evaluated the performance of the novel AI-based language generator ChatGPT in glioma-related treatment recommendations. ChatGPT correctly identified the cases as CNS tumours but lacked precision on tumour subtype. The treatment strategy and regimen recommendations were rated as good; however, it lacked the ability to nuance its recommendations when taking into consideration the functional status. Overall, our findings suggest that ChatGPT has potential as an adjunct to the multidisciplinary TB decision workflow, provided that further algorithmic advancements are made in the medical domain.

## Data Availability

All data produced in the present study are available upon reasonable request to the authors.

